# A triangulated study on non-pharmacological management of Alzheimer’s disease in Mauritius

**DOI:** 10.1101/2021.04.27.21255997

**Authors:** Geeta Devi Dorkhy, Goorah Smita, Sorefan Ameenah

## Abstract

Dementia is a neurodegenerative disease, with more than 50 million people worldwide. Nearly 60% are living in the low and middle-income countries [1]. Mauritius has a rising population of elderly people, of 7.5% above 65 years [2]. Currently 14,000 people with Alzheimer’s disease (AD), show very modest improvements with pharmacological therapies. Alzheimer’s Association in Mauritius, the only NGO, which assist person with AD and offer multiple non-pharmacological therapies (NPT).

**Objectives:** The main study aim is to find the outcomes of NPT in the management of AD and the correlation of data in mixed studies.

**Setting:** The participants were recruited from Alzheimer’s Association Mauritius, the only NGO. The center provides NPT such as cognitive training, reminiscence therapy, socialization and group interactive games.

**Primary and secondary outcome measures:** Both quantitative and qualitative studies were triangulated to find similarities (matched) and unsimilar (unmatched) results.

**Result:** Quantitative data (n=42) analysis showed a direct statistical decline in higher function (IADL) with respect to decreasing cognition. Qualitative study (n=20) emerged with 7 main themes among which ‘dependency in AD’ was a main theme. Mixed study results showed 6 out of 7 main themes were linked to/ matching to statistical results after triangulation.

**Conclusion:** NPT is an effective tool in the management of AD in Mauritius. Triangulated studies allowed in depth analysis of the patient.

**Strengths and limitations of the study:**

- Mixed study provides in depth analysis
- Data speaks for itself, it is lived experience
- Mixed studies complement each other
- However, it is a time consuming process
- Participant unwillingness to participate

## Introduction

Dementia is a broader term used in neurology while Alzheimer’s disease (AD) is a major form of dementia. In today’s world, there are about 46.8 million people living with the disease [3]. This number is expected to rise rapidly if no action is taken at present. The World Alzheimer Report (2015) has projected a value of 131.5 million, a massive rise, by the year 2050. AD causes significant detriment to quality of life for the affected patient with increased burden of care for caregivers [4]. As a result, there has been major concern for research on NPT [5]. Recent studies on non-pharmacological interventions (NPIVs) may help improve the cognition, communication, quality of life (DALY) of individual with AD [6]. More complex activities (IADL) are lost early in the course of AD, and contribute to the (DALY) disability associated with the AD [7]. So far, no research has been done on the Alzheimer patient in Mauritius, little is known on the NPT. Therefore, to be able to better understand the current situation this project was conducted at the NGO, at Belle Rose in Mauritius.

## Materials and methods

### a. Design of mixed studies

“Triangulation is above all a state of mind, which requires creativity from the researcher…” (Decrop, 2011). Borkan, (2004) stated that mixed methods or multimethod techniques is “the power that mixed methods and multimethod research possess”. As Creswell et al noted, “This form of research is more than simply collecting both quantitative and qualitative data; it indicates that data will be integrated, related, or mixed at some stage of the research process.” The integration of combining both quantitative and qualitative produces a sum greater than individual study.

The study design is different for each study however, the same pool of patients were recruited. Quantitative study started with data collection from questionnaires. Pre-validated questionnaires such as the Mini Mental State Examination, (MMSE), Activities of Daily living (ADL) and Instrumental Activities of daily Living (IADL), were used and analyzed statistically using SPSS software and chi-square tests.

On the other hand, thematic analysis was used in qualitative interview analysis. The third step was to combine the 2 methods thereafter analyzed using triangulation. Using linking process to identified and then compare to theoretical background. Triangulated data was interpreted as “matched results” or “unmatched results” and interpretation was carried out. **Figure 1**, depicts the research design.

**Figure 1:**
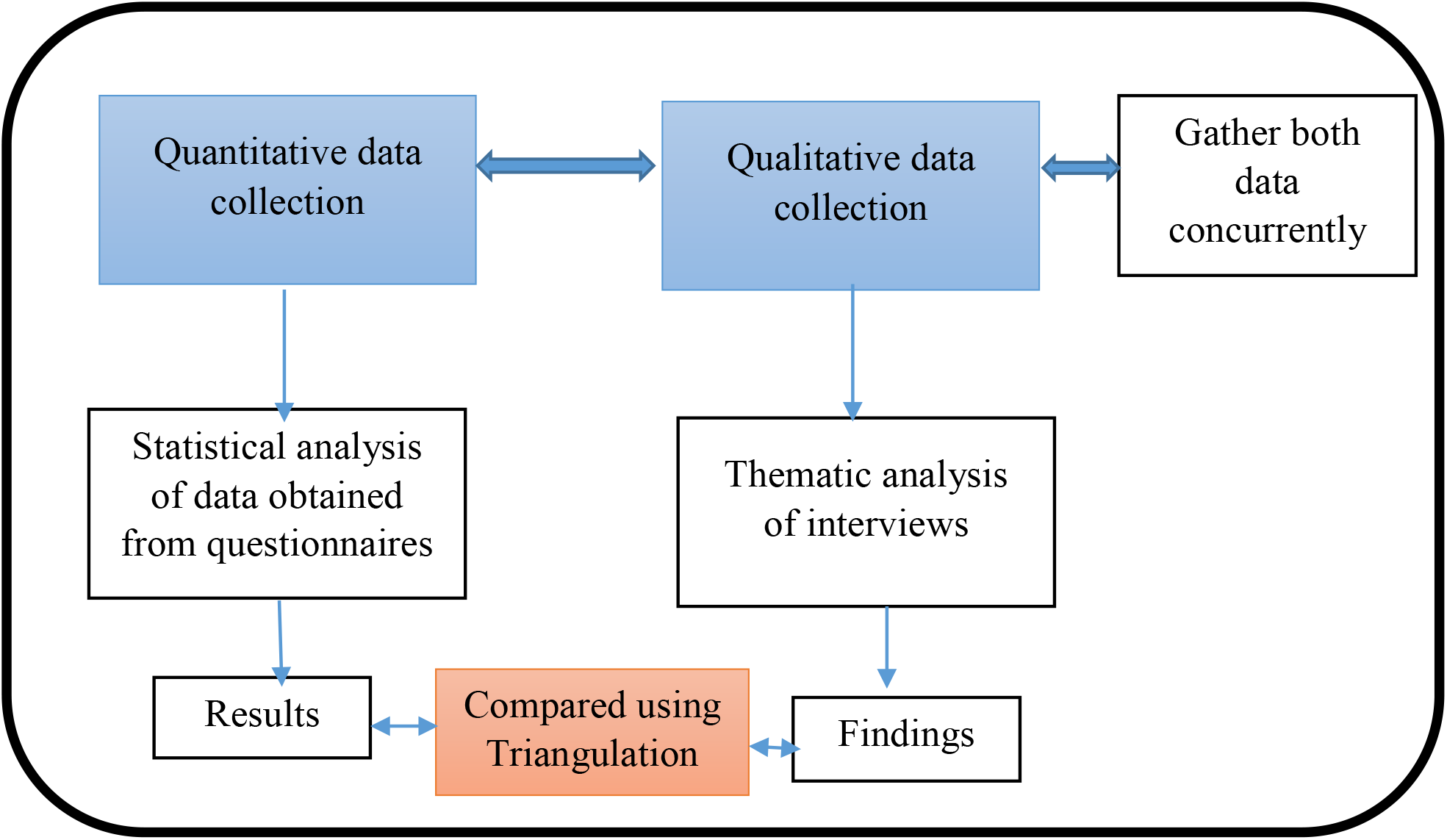
Methodological triangulation showing research design in quantitative, qualitative and mixed methods approaches (Adapted from: Twycross, 2004)

Information about the study was provided to all participants at the Association Alzheimer and subsequently invited to participate. Participation was on voluntary basis and they could leave at any point of time during the study. Information sheet was also provided and relevant details of the project was provided to the patients as well patient’s carer prior to the start.

Consent was obtained through the care giver and the patient. The process and the aim of the study was explained in simple Creole/French language. Verbal consent from both parties (carer and patient) and written consent was obtained from the carer/ care giver and /or the patient

### b. Methodology in quantitative study

#### i. Study design and process

The design of the study is quantitative research method through questionnaires and surveys. The data collected is statistically analyzed then computed using statistical techniques to find association between the variables. The results were correlated in the form of numbers and finding associations (Earl, 2010).

#### ii. Measuring instruments and procedures

Demographic details including personal information regarding name, age, gender, address, the next of kin, the medical history, past medical and surgical history, the personal history. Pre-validated questionnaires such as the Mini Mental State Examination, (MMSE), Activities of Daily living (ADL) and Instrumental Activities of daily Living (IADL), were used.

#### iii. Study numbers and sampling

A representative sample is required for conducting the survey

#### iv. Sample size calculation by Stoven formula

Sample Size: From the ‘Association Alzheimer’, 100 patients have been diagnosed with Alzheimer’s disease at the ‘Association Alzheimer’. With a margin of error of 5%, and confidence interval 95%, and population size of 100, the sample size is given by **Figure 2**.

**Figure 2:**
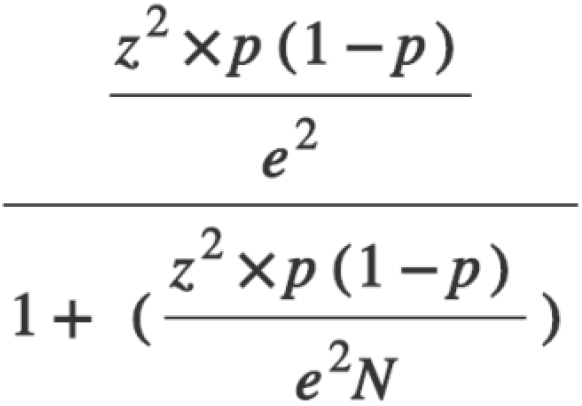
Stoven Formula used for sample size calculation

For a confidence level of 95%, α is 0.05 and the critical value is 1.96. Then, using the above formula, the calculated sample size is 80.

#### v. Inclusion and exclusion criteria

All the participants were recruited from the ‘Alzheimer’s Association’ Centre at Belle Rose, Mauritius. The elderly population aged ≥60years were included. Mild to moderate cognitive impairment and person with AD were recruited. Common local language used was Creole, Hindi, French and English. Other neuropsychiatric disease other than AD was the only exclusion criteria.

#### vi. Participation rate

The participation rate was only (n=42) in this study, there was no duplication of data.

#### vii. Data collection

The interaction between the researcher and the patient took place in a consultation room around a table, at the Alzheimer’s Association, Belle Rose, Mauritius. Prior consent obtained from patient and the carer/guardian before start. The questionnaire (MMSE) was filled in by the interviewer. Also, the ADL and IADL were used for data collection, each lasting 15 minutes. The activities included was from the MMSE-questionnaire either French or Creole was common language used among the patients at the Centre, and often English translation was required.

#### viii. Use of software

SPSS, Microsoft Excel was used for statistical analysis in quantitative study.

#### ix. Reliability

Check for reference and look citation searches in PubMed and Google Scholar.

#### x. Validity

Medical records were cross checked for demographics and past and current medical history as well as non-pharmacological therapy and progress.

#### xi. Methods and test used in data analysis

Chi square test for independence, inferential analysis, multiple regression analysis.

### c. Methodology in qualitative study

#### i. Measuring instruments and procedure

Interviews were carried out in qualitative design (study 2) using open ended questions as shown in.

#### ii. Methods and materials

Qualitative research is to understand the people from their own perspectives and their own frames of reference, how they live their daily lives, experiencing reality.

#### iii. Selection and description of participants

All participants were recruited from the Alzheimer’s Association, at Belle Rose, Mauritius. Sample size was obtained till theoretical saturation reached and no new ideas, data emerged. Interview questions were open ended and clear consisting of 20 open-ended questions. Some instructions were provided for exploring more details, and to metamorphose between topics. The interviewer proceeded to the interview process by engaging the patient in an initial discussion of what are the main types of NPT and how much they were involved.

All interviews were audio recorded and then later transcribed. Person living with Alzheimer’s disease own experiences was written down, noted and recorded with the use of smartphone and later translated from creole or French language to English using google translation. Spoken words and observable behaviour of the individuals formed part in developing a concept.

Eventually these were grouped together as particular concepts/themes which generated the field notes.

Inclusion and exclusion criteria same as above

#### iv. Technical information

Phenomenological interpretative analysis method in qualitative research is an ongoing process of coding data and decoding and grouping similar chunk of data together.

The following steps are required: Collecting data (Creole language), proceed to coding of data, followed by “categorizing bits of data and then interpreting interview data and writing field notes” [8]. “Initial coding involved describing and labelling units of data”. Also “similar kinds of information are grouped together into categories”. Then “relating different ideas and themes to one another” [9]. This process was repeated until all raw data are coded once.

This step also involves literature reading. Together with the “naïve understanding and the authors’ pre-understanding, these were reflected by using suitable literature to form a comprehensive understanding” [10]. Lastly interpretation takes place.

#### v. Procedures

No software was used for data processing and analysis in qualitative study. At time of study, there was no exposure to software material for the duration of the research. The following key terms were searched across all the online databases: “Alzheimer’s Disease”, “Dementia”, “lived-experience”, “dependency”, “emotions”, “quality of life”, “physical exercise”, “group singing”, “music playing”, “family and carer”, “anxiety”, “decision-making”, “interactions with others”, “having a pet”, “outdoor activities”, “awareness into illness”. Main themes and sub-themes emerged in qualitative research.

#### vi. Theoretical saturation

A relatively small number of respondents (15-20) until ‘theoretical saturation’ when little new comes out of additional interviews [11].

#### vii. Use of software

No software was used for data processing and analysis in qualitative study 2. **Figure 3**, differentiates between quantitative and qualitative study in table form below.

**Figure 3:**
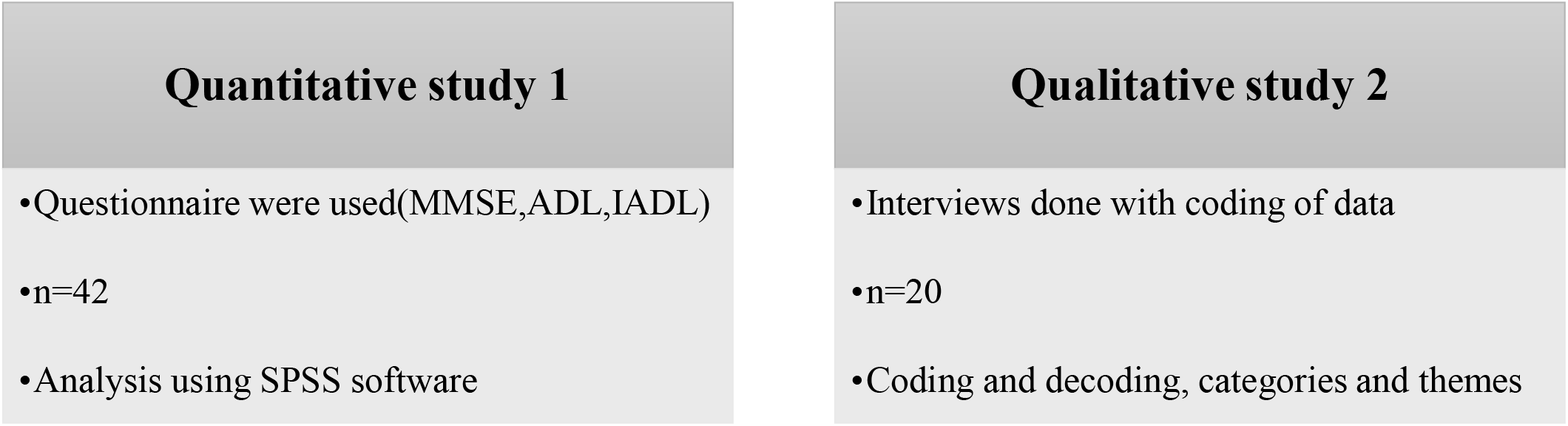
Quantitative study v/s Qualitative study

### d. Methodology in triangulated study

Triangulation means using more than one method to collect data. Also, Patton, (1999) added that triangulation is a comprehensive understanding of phenomena while “method triangulation involves the use of multiple methods of data collection about the same phenomenon” [12].

In this study, both quantitative and qualitative study has been combined (triangulated) as depicted in **Figure 4**. Therefore, it is an integrated research study method.

**Figure 4:**
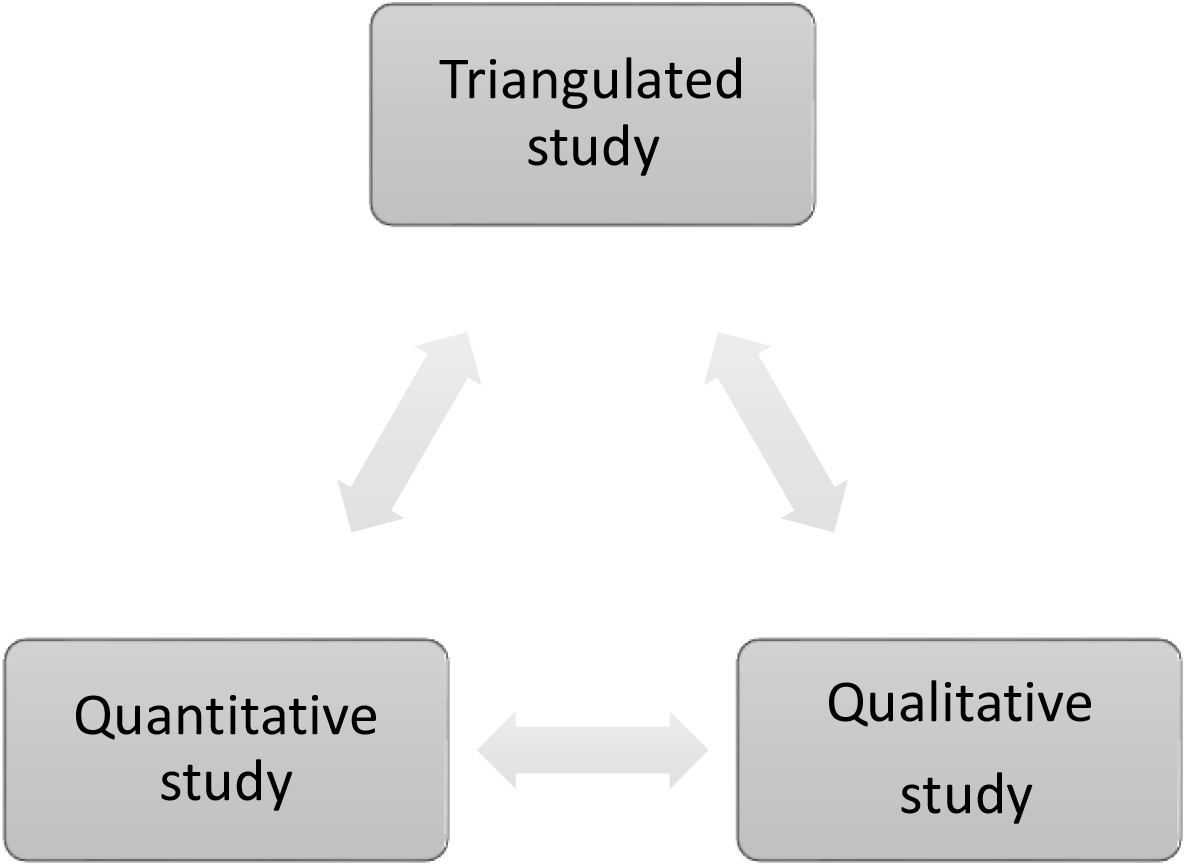
Mixed method study design in triangulated study

### e. Patient and public involvement

The main research topic was revolving around the NPT of AD in Mauritius. The patients were recruited from Alzheimer’s Association Mauritius, the only NGO. They were already on NPT. They were involved in the quantitative and qualitative study design and recruitment to and conduct of the study. The results of the study will be disseminated to study participants during annual parent group meeting.

### Analysis and findings in triangulated study

Matching has been done with combination of statistical data and themes from each individual studies. This has been tabulated (**Table 1)**. The interpretation of the triangulated data has also been given in last column.

**Table 1:** Analysis (of mixed studies) in triangulated study.

| Qualitative study |  | Quantitative study |  | Triangulated study |  |
| --- | --- | --- | --- | --- | --- |
| Scale | Statistical Analysis | Main theme | Sub-theme | Status (Matched/unmatched) | Interpretation |
| Functional capacity<br>IADL (Score:0-7)<br>ADL (Dependent/independent) | IADL score of 7 indicates an independent function (frequency 1)<br>IADL score 6,4 equal to frequency distribution of 9 | Activity | Exercise (stretch, yoga, walking),<br>breathing exercise, throw ball | Matched result | ADL is the most basic self-care<br>IADL are the slightly more complex skills<br>Activities undertaken by person with AD have functional capacity |
| Cognitive capacity<br>MMSE | Normal cognition<br>53.7% | Music | Like singing, Play instrument | Matched result | Normal cognition in person with AD show ability to sing, play instrument |
| Cognition, MMSE (orientation to time, place and person, memory, language, registration) | Mild cognitive impairment 19.5%<br>Severe cognitive impairment 26.8% | Music, singing | Repeated same words, sentence<br>Forget, mix up<br>Poor recall<br>Language - difficult, incorrect words, long pause | Matched result | Mild, severe cognitive impairment affects the brain functioning in AD |
|  |  | Caring | Care for pets, dog, birds | Unmatched result |  |
| MMSE, Cognition | Mild to severe cognitive impairment (19.5%, 26.8%) | Knowledge on AD | Lost key, wallet associated with the disease | Matched result | Higher function like recall is affected in AD |
| MMSE on ADL and IADL | MMSE with ADL (p-value >5%)<br>MMSE with IADL (p-value : 0.006) | Dependency | Travelling, spending money, Doing house chores, Cooking | Matched result | Higher complex functions in IADL is affected with decrease in cognition, so does the level of |
| ADL | No functional impairment in 73.8%<br>Mild and severe functional impairment (14.3%, 11.9%) | Autonomy |  |  | Dependency in AD in some of the patients. |
| ADL |  | Family | Support, caring, friends, happiness | Matched result | Family support is important, as patients need help with activities of daily living |
| MMSE | Normal<br>Cognition in<br>53.7%, mild and<br>severe<br>impairment<br>(19.5%, 26.8%) | Recall<br>memory | Past, recall,<br>forget | Matched<br>result | As the cognitive<br>function declines, the<br>recall potential<br>decreases in AD |

**It is important to note that both matched and unmatched data in triangulation is of great importance in analysis and data interpretation, Figure 5**.

**Figure 5:**
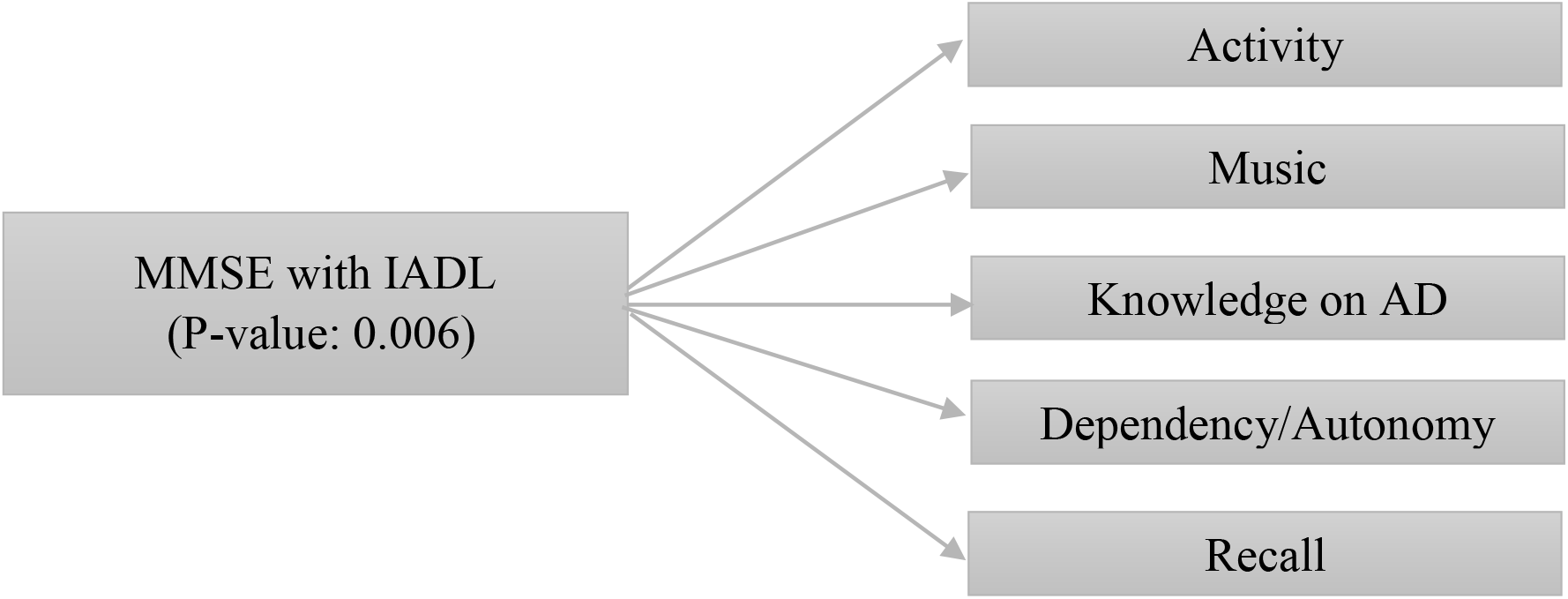
Summary flowchart linking quantitative data (study 1) with qualitative themes (study 2). Multiple correlation analysis in study 1, showing matching in different aspects/ themes in study 2.

MMSE with IADL (p-value: 0.006) showing that a decrease in the complex functional status (IADL) resulting in decrease with cognition (decrease MMSE) in AD. (Quantitative study 1). The same has been reported during the interviews in qualitative study 2. The functional status and level of activity, the cognition, knowledge, level of dependency, autonomy and recall memory of the person with AD.

## Discussion in triangulated study

The assessment of older people is a complex process requiring evaluation from different aspects. The aim of this study is to compare the different management strategies and the outcomes of the person with AD. Results varied with different level of dependency and autonomy in various aspects of living with the disease. Quantitative research resulted in numbers while qualitative study captured the lived-experiences and emotions of the patients with AD with respect to NPT.

AD is a complex disease and therefore the management of this group of individuals is also complicated. Triangulation is a challenging task since it involves the integration of data and findings [13]. Triangulated data analysis complemented each other in various dimensions of the disease aspect, especially since AD is a progressive and irreversible brain disorder. These patients have to be treated with dignity and respect. Both positive and negative findings were explained.

Most of the sampled population were in the age group of 81-85 years (highest frequency of 10) while there were more females than males group (30:12). Their duration of stay varied from 1-5 years at the center. In terms of numbers though above 50% of the population showed normal cognition as well as normal ADL it was important to consider the other half with varying results as discussed below. AD is classified as mild, moderate and severe cognitive impairment based on MMSE. 53.7% AD patients showed normal levels of cognition, while 19.5% had mild and 26.8% with severe impairment. Not being able to perform well in cognitive functions represented partly this 19.5% and 26.8% for mild and severe loss in cognition.

73.8% of the study population represented the independent states on ADL scores. This was also linked to the fact that in qualitative data themes emerged such as “autonomy” and “dependent” describing the various levels in performing ADL. Severe ADL was found at 11.9%, while moderate dependency was at 14.3% meaning that the patients with AD were dependent on carer or family member. The IADL component assesses more complex activities. IADL best score of 7 was represented by a low percentage of 2.4 meaning that the patients with AD were mostly dependent and this is represented by a 59.5% which indicated moderate dependency of care. This was mainly seen in the patients who could be assisted in travelling, in cleaning the house, preparing the meals, shopping, assisted in giving medicines. Assistance such as ‘a maid’ or ‘a driver’ was required, as stated. Patients mentioned ‘their son or daughter’ took care of them. Hence, showing need for care and love.

Previous studies showed that the cognition improved immediately from previous studies [14, 15]. Multidimensional stimulation therapy study also showed positive influence on cognition [16]. Results of my research showed that 53.7% of normal cognition, 19.5% mild cognition and 26.8% with severe cognition for the patients already on NPT. In addition, there was a positive correlational finding between the cognition and the complex activities of IADL, meaning that the reduced cognitive power resulted in decrease in functional activities of IADl. In mild cognitive impairment (MCI) individuals, there is no effect of exercise as reported by Gates (2011) [17]. This in contrast to another study by Sherman (2017), who showed greater efficacy in multi-modal strategies [18]. Similarly, there has been improved cognition and function in PWD with both pharmacological and NPT [19].

No correlation was found between the variable and cognition in my research as mentioned above. Similar findings from Conde-Sala et al. (2012), who reported no relation between variables such as age, gender, schooling level with MCI-AD after a five-year follow-up [20]. This was in contrast to Meguro K et al. (2007), who analyzed and concluded variables have significant effect on the progression of AD [21].

Further finding of the age factor with cognition in a report showed that there were 0.5 million centenarians in the world (2016), accounting for approximately 7.5% of people aged ≥65 years. Thus, dementia seems to suggest that it is not a consequence of extreme old age. However, Paul Denver (2018) reported that advanced age was a major risk for but sub-clinical cases are difficult to identify until very late [22]. This was not found in my study as there was no relation between cognition and age factor.

Dementia and AD potentially affect more women than men (Mauritius statistics, 2014) [23] Women are at higher risk of developing AD than men (Rita Guerreiro, 2015) [24].

Other risk factors such as type 2 diabetes mellitus and high blood pressure, also contribute to AD, as Lontchi-Yimagou et al (2013), concluded in his study as inflammation is common for the setting of AD [25]. But there were no such findings in my research.

Data also suggest the higher the level of education, the better the neuronal coping abilities in later life as displayed in a study [26].

My research findings show no correlation between smoking and cognition in AD patients. In contrast to Hisayama Study, which was a large cohort study in, Japan, included PWD aged 65 to 84 and they were followed for 17 years from year 1988-2005. He concluded that 252 subjects, on follow up, developed dementia, while 143 persons developed Alzheimer’s disease. It was also noted that smoking increases risk of dementia.

Zhong G et al. (2015) assessed the risk between AD and vascular dementia associated with smoking using online database and concluded that there was an increased risk of dementia is associated with smoking [27].

A positive correlation between IADL and MMSE was found in my research. The assessment of AD is important, the more the decline in the cognition or function of the patient with Alzheimer’s disease, the worst the outcome [28]. AD manifests itself as reduced cognition and reduced activities of daily living (ADL) (Saari T, 2018). The brain is a complex structure and higher functions are performed by the brain. Since the activities are coupled with each other, therefore the ability to recognize a stimulus is complex and requires previous exposure and recall. These are also influenced by multiple factors which is difficult to understand separately (Eichenbaum et al., 2019) [29].

### Strengths of triangulated study

The richness and variation of the data show “quality is more important than quantity when it comes to examinations of the life world” (Dahlberg, Dahlberg, & Nyström, 2008). One research study method can complement the other study method and produce better results and interpretations [30].

### Weaknesses and limitations of triangulated study

Factors which may have influenced the results and also present as limitations in the study are discussed below. Due to old age, many patients were having hearing problems, decreased vision and lack of cooperation and difficulty in understanding instructions. It is a time consuming and tiring process for the patients to be able to interact with the researcher during the whole process of data collection. Low educational background, their reading and writing was minimal, and English was difficult to write and read. Unwillingness to participate further. Assessment of older people is a complex process requiring patience and time. It was difficult to obtain them for voluntary participation. Also, some patients were reluctant to fully participate for the whole duration of the study process. Findings can be mixed up in triangulation at the costs of different methods used and quality of study. It is expensive also and time consuming. Lack of prior research studies since very little is known on AD in Mauritius, since no research has been conducted specifically in the non-pharmacological management of this group of people.

## Conclusion

Consequently, the results obtained in triangulated study would contribute to add to further knowledge and also to help plan and devise more appropriated, adapted treatment for the aging Mauritian population. In the near future, more research, involving a larger population sample and a more dynamic and robust study design aiming at the management strategies would emerge to prevention and management of the person with higher risk of AD.

## Data Availability

All data available in the manuscript may be provided upon request

https://www.alzheimerdisease.com

https://www.alzheiemrmanagement.com

## Contributorship statement

The research has been possible due to two important persons. During the planning process Dr Ameenah Sorefan and Dr Smita Goorah and myself, Dr Dorkhy Geeta Devi, we were involved in planning the project work. Important points were analyzed in depth during the conception and design, acquisition of data from the pool of patient. During the next step of research, both Dr Smita Goorah and myself, Dr Dorkhy Geeta Devi were actively involved for conducting and reporting of the work. Data analysis and interpretation was also contributed by both of us.

## Data sharing

No additional data available

## Acknowledgement

Thanking all the parents, carers, persons with AD and Mr Raj Guness, those who helped made this research possible. I am grateful to my teacher Dr Suranjana Ray, Dr Namrata Chhabra and Dr Ranjit Patil, from SSR Medical College for their constant guidance. The Lord Almighty and my parents have unconditionally provided support and their blessings at each turning point of life.

## Ethics statement

Ethical committee the Ministry of Health and Quality Of Life, Mauritius and the Consultant, Ministry of Social Security, National Solidarity & Reforms Institutions, Mauritius (ID: MHC/CT/NRTH/GEEDO).

